# Medical Students’ Knowledge, Attitudes, and Motivation towards Antimicrobial Resistance Efforts in Eastern Uganda

**DOI:** 10.1101/2024.11.09.24317028

**Authors:** Jonathan Babuya, Daniel Waruingi, Douglas Mungujakisa, Osmas Ahimbisibwe, Victoria Ruth Kako, Faith Aporu, Emmanuel Mugume, Julian Nyamupachitu, Kenedy Kiyimba

## Abstract

**Introduction:** To promote holistic learning about Antimicrobial Resistance (AMR), catalyze multidisciplinary engagement, and innovative AMR interventions, it is important that learning goes beyond the classroom and students embrace different extracurricular interventions. This study aimed to determine knowledge, attitudes, and motivations influencing medical students’ engagement in AMR Club Initiatives at Busitema University in Uganda.

**Methodology:** This was descriptive cross-sectional study conducted at Busitema University among undergraduate student pursuing Bachelors of Medicine and Surgery, Bachelor of Nursing, and Bachelor of Science in Anesthesia and Critical care. A semi structured pre-tested questionnaire was shared among the study participants. Bloom cut-off method was used to analyse the knowledge of the participants, chi square test used for bivariate analysis and multivariable logistic regression used for determining factors independently associated to students’ engagement in AMR club activities.

**Results:** 71.5% of the 193 students had sufficient knowledge (determined using Bloom’s cutoff categories from 60% and above as sufficient and below 60% as insufficient) on AMR with an average score of 68.18% (SD= ±16.12). 90% of participants recognized the significance of incorporating AMR within their curriculum, and 87.5% appreciated the need for training AMR using a One Health ApproachThe most common reasons why students engaged in extracurricular efforts to address AMR such as forming AMR clubs were found to be; influence from peers (n= 42), university support (n=35), and inspiration from peer mentors’ work (n=35).

**Conclusion:** The students were found to have a high level of knowledge and positive attitudes towards AMR but reported the need for further in-depth training. Extra-curricular engagement such as participation in an AMR Club was found to positively influence students’ engagement in AMR interventions.

## Introduction

Antimicrobial Resistance (AMR) has been declared by the World Health Organization as one of the top 10 global health threats[1]. Despite the high mortality and burden caused by AMR, it is largely ignored and rarely prioritized relative to other global health threats[2]. In 2019, 4.95 million deaths were associated with AMR globally, and 1.27million deaths were caused directly by resistant infections[3]. In Sub-Saharan Africa, bacterial AMR was associated with 1.05 million deaths and attributed to 250,000 deaths[4]. In Uganda, 7,100 deaths were directly caused by bacterial drug-resistant infections, and 30,700 deaths were associated with drug-resistant infections[4].

Education and awareness play a very critical role in antimicrobial use and shape other health-seeking behaviors that influence AMR[5]. The Global Action Plan on AMR developed by the World Health Organization (WHO) lists improving awareness and understanding of AMR as its first objective[6]. It highlights the need to raise awareness and promote behavioral change through various ways such as public communication programs implemented from a One Health realm targeting human health, consumers, animal health, and agricultural practices. It also makes a case for inclusion of AMR into the curricula, and the professional training of health, veterinary, and agriculture sectors to promote a better understanding among professionals. The Uganda National Action Plan on AMR strategic objective one proposes the promotion of public awareness, training, and education through two major strategies; improving public awareness and supporting education and training of human, animal, plant, and environmental health through both pre-service and in-service training. Educating them very early in their training and practice provides them with a different perspective enabling them to challenge existing norms that undermine antimicrobial stewardship and also serve as AMR champions in their respective facilities. There have been several efforts in Uganda to incorporate AMR into the curriculum but they are yet to be successful[7].

A study conducted across East African Universities, reported that AMR was not emphasized in the medical curricula[8]. Comparable findings were reported in Ethiopia, Ghana and Zambia among nurses and pharmacy personnel and healthcare students’ in regard to knowledge of Antibiotic use, AMR, and antimicrobial stewardship programs[9]. In Uganda, while the medical students were aware of the concept of AMR, they lacked in-depth knowledge necessary for effective antimicrobial stewardship practices[10]. A research conducted amongst medical, pharmacy and nursing interns also showcased that only a small percentage of participants had good knowledge of AMR and rational prescription practices[11].

To promote behavior change, and inspire action among healthcare students, engagement should extend beyond traditional classroom settings to include holistic learning that incorporates both curricular and extracurricular activities[12]. From early in their training, tertiary level students should be empowered to engage in AMR. Extracurricular activities such as participation in clubs have been shown to enhance intrinsic motivation, and active participation in efforts to address various global health challenges[13]. To develop supportive frameworks where healthcare students can engage actively and empower communities on AMR, it is best to understand their perspective and drive to engage in such initiatives[14].

Currently, there is a paucity of data from African tertiary institutions on engagement of healthcare students in extracurricular activities on AMR. It is important to have evidence which can be harnessed to promote effective and beneficial extracurricular engagement among healthcare students. This study aimed to determine knowledge and perceptions on AMR among healthcare students and the motivation that influence healthcare students’ participation in AMR extracurricular activities such as clubs.

## Methods and Materials

### Study Design

This study was a descriptive cross-sectional study employing quantitative data collection method to assess the level of awareness and knowledge of antimicrobial resistance and factors influencing healthcare students’ engagement in AMR club initiatives at Busitema University.

### Study setting

The study was conducted at Busitema university, Faculty of Health Sciences in Mbale, Eastern Uganda between December, 2023 and February, 2024.

### Study population

These were undergraduate medical students pursuing Bachelor of Medicine and Bachelor of Surgery, Bachelor of Science in Nursing, Bachelor of Science in Anesthesia and Critical care at Busitema University.

### Eligibility criteria

The study included undergraduate students at Busitema University, Faculty of Health Sciences who were 18 years of age or older pursuing either Bachelor of Medicine and Bachelor of Surgery, or Bachelor of Science in Anesthesia and Critical Care, or Bachelor of Nursing Science. The students who were not at the Faculty due to other academic programs, such as anesthesia rotations or in their last semester of study were excluded.

### Study variables. Dependent variable

The dependent variable was students’ engagement in AMR related activities.

### Independent variables

These included; socio-demographic characteristics like the age, sex, year of study and course of study, AMR Awareness, Knowledge levels on AMR, and Extracurricular activities engagement.

### Data collection tool and procedures

We used a semi structured questionnaire which was self-developed and pretested before use. We uploaded the questionnaire onto KoBo Toolbox. KoBo Toolbox is an open-source software developed by the Harvard Humanitarian Initiative with support from United Nations agencies, CISCO, and partners to support data management by researchers and humanitarian organizations (https://www.kobotoolbox.org/). The servers are secure and encrypted with strong safe guards and protection against data loss. Participants who consented to the study had the questionnaire link sent to them via a WhatsApp message. The data collection tools were pretested before actual data collection.

### Sample size estimation and sampling technique

The sample size was determined using the formula by Fisher et al. (1998): n= (Z2 x p(1-p))/d2 Where: n= required sample size, *Z* = standard normal deviation at a 95% confidence interval (1.96), *p* = prevalence (0.5 due to no similar study found), *d* = margin of error or degree of accuracy (0.05) After calculations, *n*=384.16, rounded to 385 participants. Since the population was less than 10,000, the finite population correction formula was applied. With a population size of 500 students, the final sample size was determined to be 218 study participants.

A list of all the students at the faculty was obtained, grouped as per the year of study. Random numbers were assigned to the names, they were then filled in a random selector program, that generated a random list of the numbers to which the names were attached. The selected students were then approached via a phone call or WhatsApp and given information about the study. Those that consented to participate in the study were recruited and the link to the questionnaire was sent to them. The reasons for engagement in AMR activities were first collected through a pre-study survey among 20 students to populate the questionnaire since we could not find any qualitative study that had been done about these reasons.

### Data analysis and presentation

The data collected was meticulously cleaned and analyzed using STATA version 15.0. Categorical data was analyzed using frequencies and percentages, while numerical data was analyzed using measures of Central Tendency and Dispersion. Pie charts were used to visually represent categorical data such as gender distribution, age groups, study programs, and year of study while bar graphs used to illustrate comparisons, such as students’ engagement in extracurricular activities by gender, knowledge levels about AMR across different programs, and motivations for engaging in AMR club activities. Bivariate analysis investigated factors associated with students’ engagement in AMR activities i.e. age groups, study program, year of study, knowledge about AMR, awareness about AMR, and participation in extracurricular activities and presented in tables while multivariable analysis further examined the significant factors identified in bivariate analysis to determine their combined impact on students’ engagement in AMR activities and presented in figures including pie charts, bar graphs, and other visual aids to succinctly display key findings.

### Ethical consideration

The study was conducted according to the Declaration of Helsinki. Ethical approval was sought and obtained from Busitema University Research Ethics committee under the reference number (BUFHS-2023-104) . Written informed consent was obtained from the participants before recruitment into the study. The data collected was kept in a locked folder in the computer which was accessible only to the investigators.

## Results

### Social demographics of the participants

A total of 193 students participated in the study. The majority of respondents were male (58.0%), while females comprised 42.0% of the participants. Over half of the respondents (51.3%) were below the age of 25, 40.4% were aged between 25-34, and a small proportion (8.3%) were above 35. The largest group of respondents were enrolled in the Bachelor of Medicine and Bachelor of Surgery program (63.2%). The Bachelor of Nursing Science program had 19.7% of respondents, and the Bachelor of Science in Anesthesia and Critical Care program had 17.1%.The distribution across years of study was relatively even, with the highest percentage in Year 4 (24.4%) and the lowest in Year 2 (17.1%). Year 1, Year 3, and Year 5 had 19.7%, 20.2%, and 18.7% of respondents, respectively(**Tabel 1**).

### Students’ engagement in extracurricular activities

Extracurricular activities, in the context of this study, refer to general institutional activities outside the classroom that students participate in. The study found a difference in extracurricular engagement between genders: 82.05% of female students participated in extracurricular activities at the university, compared to 95.5% of male students (Figure 1).

**Figure 1:**
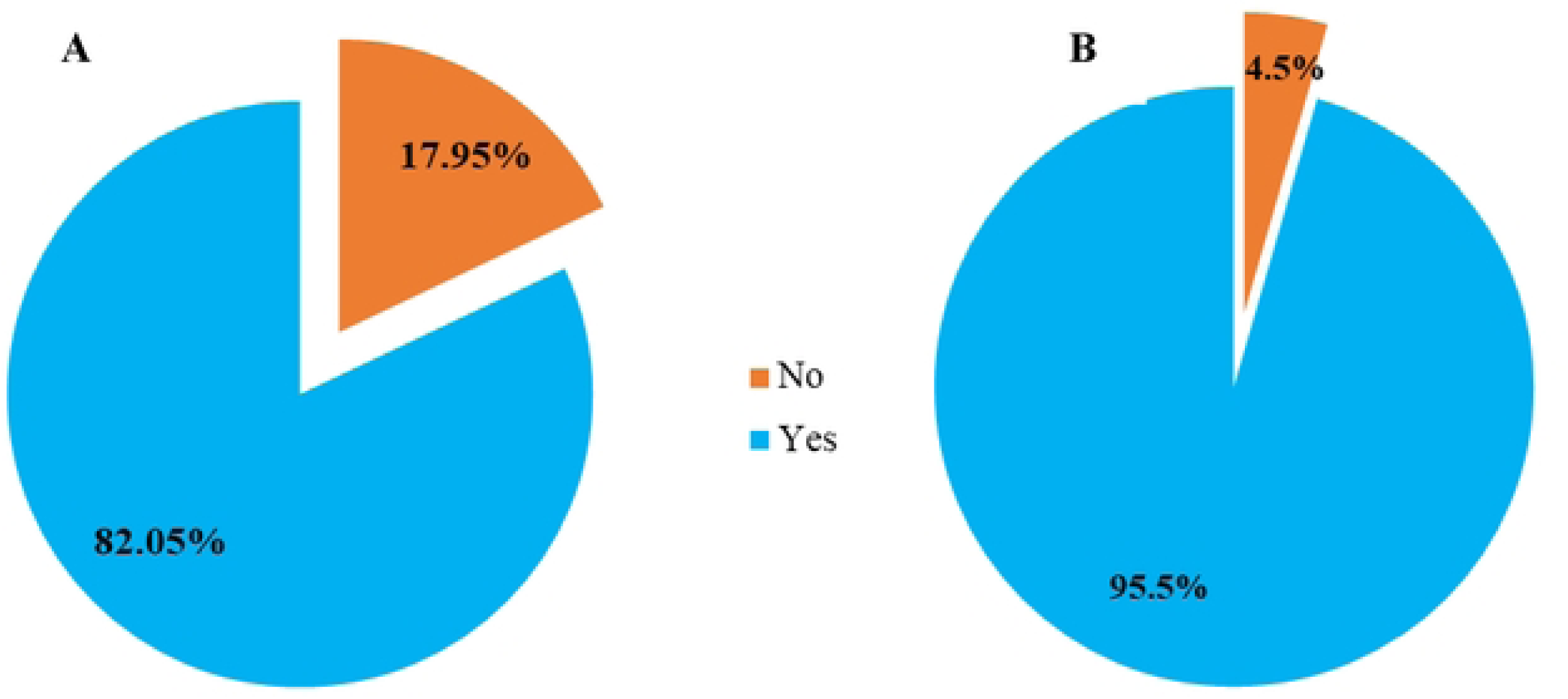
Showing students engagement in extracurricular activities; A; female, B: Male

### Students’ engagement in AMR club activities

AMR clubs, which are among extracurricular activities, focus specifically on interventions related to antimicrobial resistance. At Busitema University, 67.5% of female participants reported not having participated in AMR club activities, compared to 58.18% of male students(**Figure 2**).

**Figure 2:**
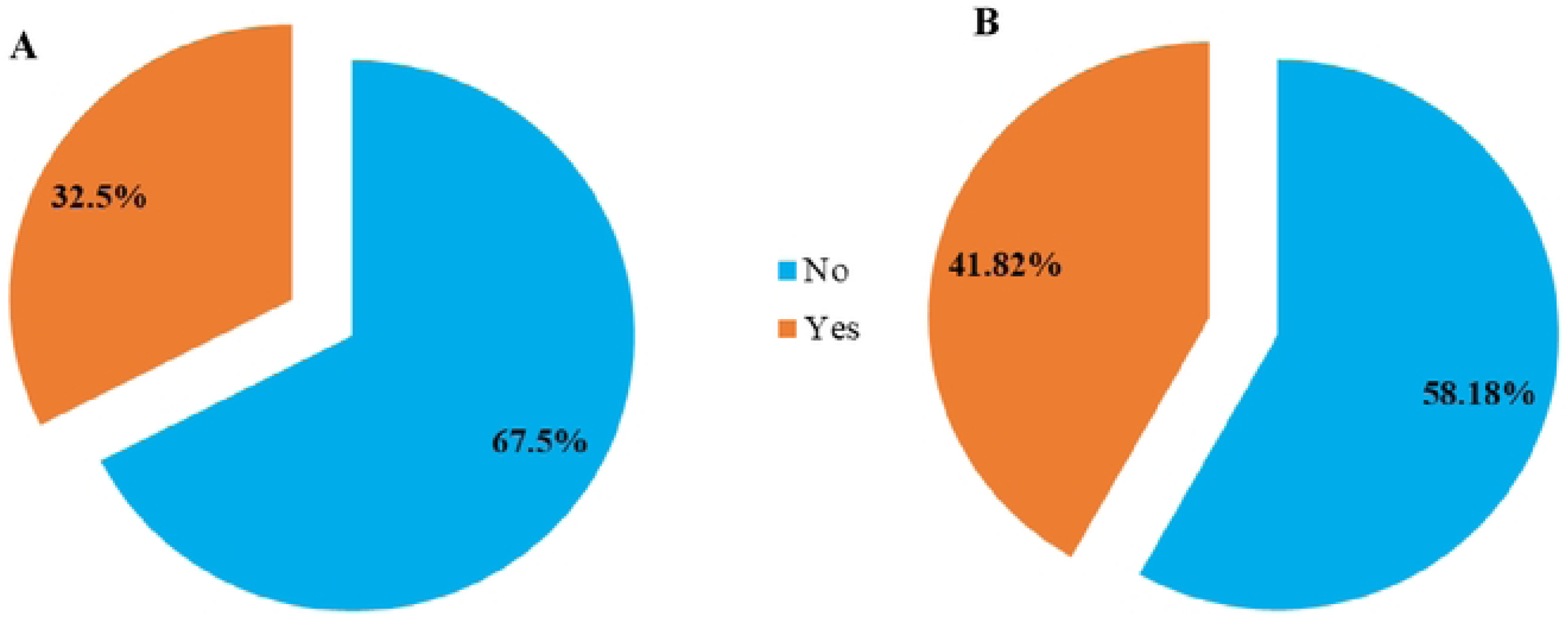
Showing students engagement in AMR activities, A: Female, B: Male

### Levels of knowledge about AMR among the students

Overall, 71.5% of the students had sufficient knowledge about AMR (determined using Bloom’s cutoff categories from 60% and above as sufficient and below 60% as insufficient). The percentage of students with sufficient AMR knowledge increased with each year of study, starting at 55.26% in Year 1 and rising to 91.67% in Year 5(Figure 3). Across the different programs of study, Bachelor of Medicine and Bachelor of Surgery had the highest percentage, 76.23% of students with sufficient knowledge about AMR which was followed by Bachelor of Science in Anesthesia and Critical Care, 69.7% and Bachelor of Nursing Science at 57.89% (**Figure 4)**.

**Figure 3.**
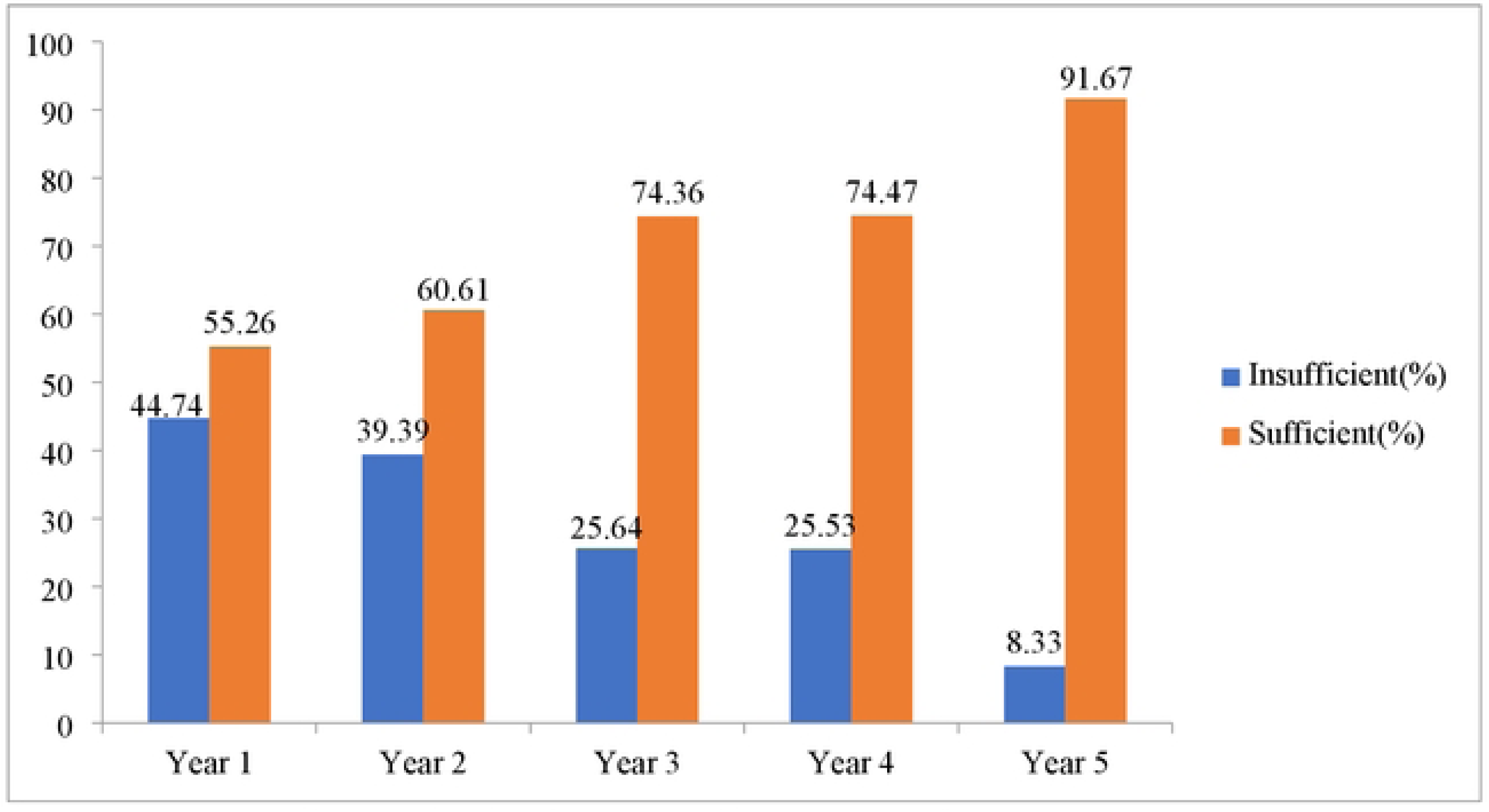
Showing knowledge levels across different years of study

**Figure 4:**
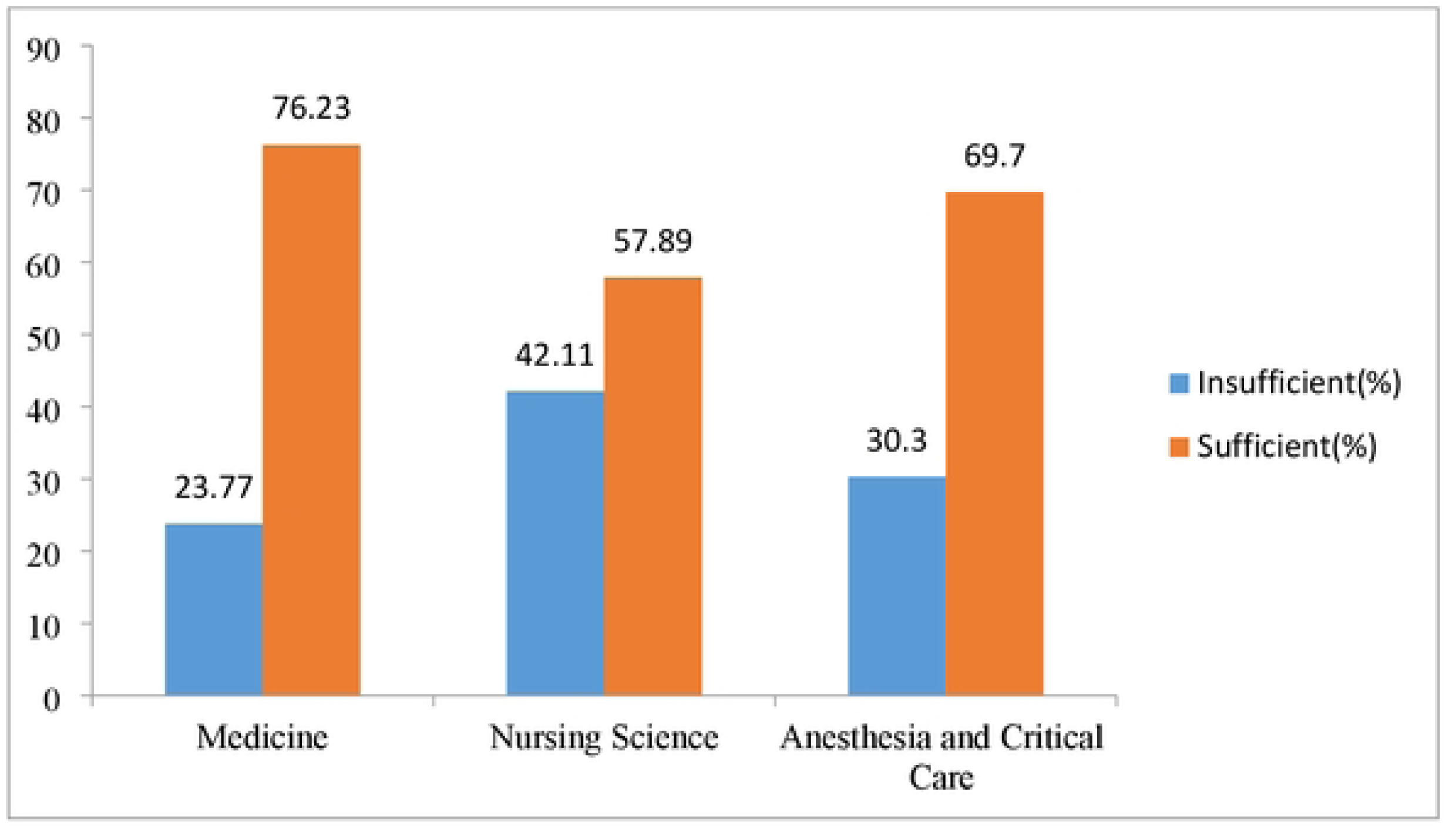
Showing knowledge levels of students in different study programs

### Attitudes of students towards AMR

A total of 8.9 % (n=17) of the study participants agreed that agreed that purchasing antibiotics without prescription from pharmacies was acceptable. Over a quarter of the participants (35.3%, n=68,) agreed to not being at risk of getting an antibiotic-resistant infection as long as they took their antibiotics correctly. Furthermore, an overwhelming majority (83.4%, n=160) agreed that prescribing broad-spectrum antimicrobials instead of narrower spectrum options contributes to antimicrobial resistance.

Majority of the particpants, 96.4% (n=186) acknowledged the critical importance of having a strong understanding of AMR in their medical careers. A substantial percentage 87.5% (n=169) agreed that poor infection control practices by healthcare professionals significantly contribute to the spread of antimicrobial resistance. Similarly, more than 80% of the students (n=164, 85%) believed that the excessive use of antimicrobials in livestock contributes to the problem of antimicrobial resistance.

Regarding their practical experiences, 73.6% (n=142) of the students reported observing instances where antimicrobials were overused during their hospital rotations. Looking towards the future, over 40% (41.4%, n=80) of the participants expressed optimism that new antimicrobial agents capable of addressing resistance issues would likely be developed. These results are summarized in **Figure 5**.

**Figure 5:**
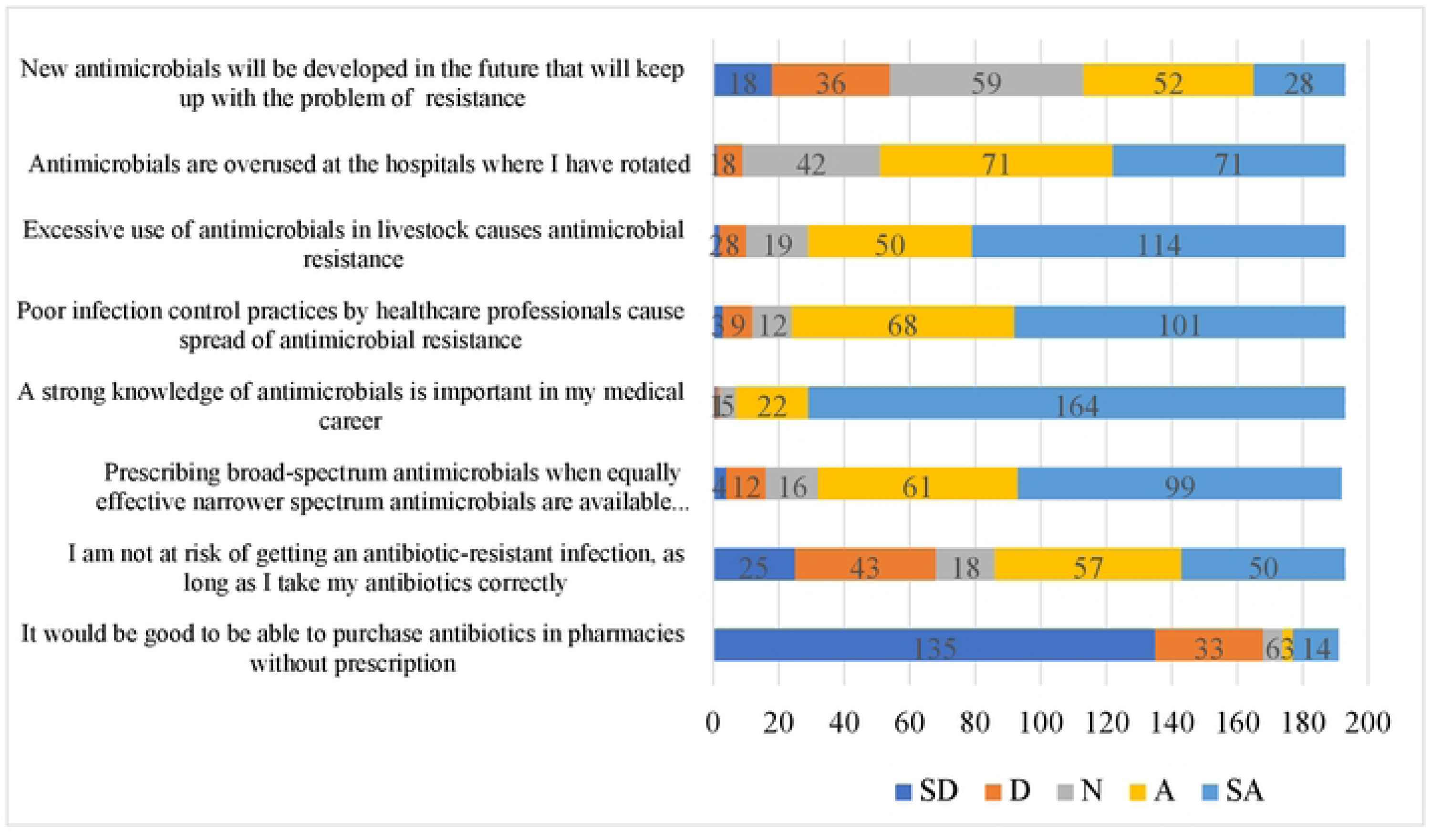
Showing students’ attitudes towards AMR: SD-Strongly disagree, D-Disagree, N- Neutral, A- Agree, SA- Strongly Agree

### Sources of information about AMR

Academic learning platforms were the major sources of information regarding AMR(n=152). (**Figure 6**).

**Figure 6:**
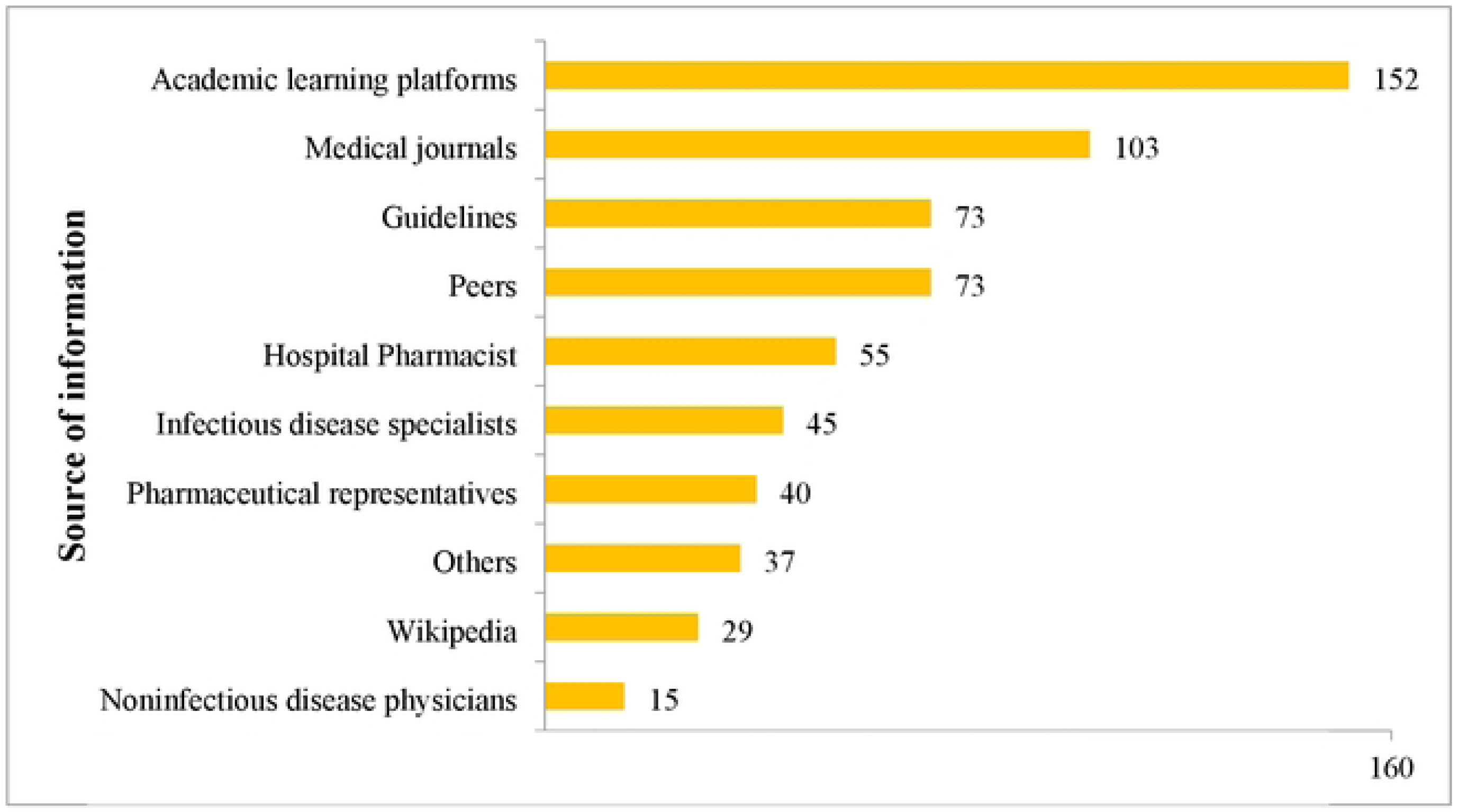
Sources of AMR information used by study participants

### Motivations to students for engaging in AMR club activities

The largest proportion of students (n=42) were those motivated by peer to engage in AMR club activities while the least number (n=4) engaged in AMR activities to get information (***Figure 7***).

**Figure 7:**
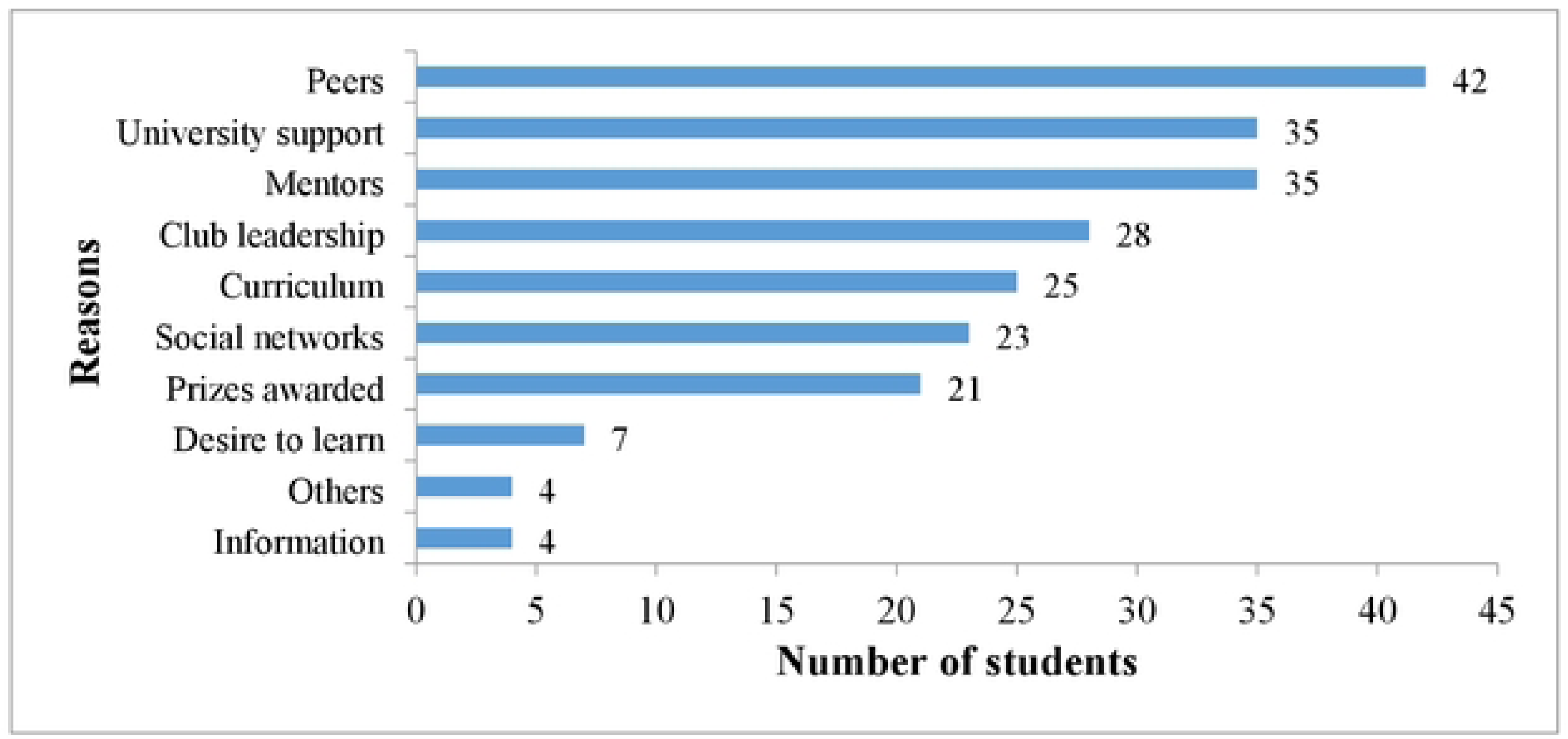
Showing reasons for student engagement in AMR activities

A higher proportion of the students (59%, n=114) students engaged in extracurricular activities like clubs to create friendships and to exercise and gain physical fitness while the least number of the students engaged in extracurricular activities due to other reasons (n=5) which included inspiring others and getting knowledge for the future (**Table 2.)**

**Table 1:** Showing the demographic statistics of the the study particpants n=193.

|  | n (%) |
| --- | --- |
| **Gender** |  |
| Female | 81 (42.0%) |
| Male | 112 (58.0%) |
| **Age** |  |
| Below 25 | 99 (51.3%) |
| 25-34 | 78 (40.4%) |
| Above 35 | 16 (8.3%) |
| **Study program** |  |
| Bachelor of Medicine and Bachelor of Surgery | 122 (63.2%) |
| Bachelor of Nursing Science | 38 (19.7%) |
| Bachelor of Science in Anesthesia and Critical Care | 33 (17.1%) |
| **Year of study** |  |
| Year 1 | 38 (19.7%) |
| Year 2 | 33 (17.1%) |
| Year 3 | 39 (20.2%) |
| Year 4 | 47 (24.4%) |
| Year 5 | 36 (18.7%) |

**Table 2:** Reasons for engaging in Extracurricular activities by the study particpants.

|  | Number of students who selected |
| --- | --- |
|  | 77 |
| To create friendships | 114 |
| To exercise and gain physical fitness | 114 |
| To develop leadership skills | 70 |
| To pass time | 51 |
| To contribute to well-being of society | 66 |
| To improve my CV | 43 |
| To get mentored into my future career | 61 |
| To improve job prospects after completing school | 41 |
| Other reasons | 5 |

### Factors associated with student participation in AMR club activities

Age, gender and program of study were not significantly associated with student participation in AMR club activities independently. Third year students had 3.12 times higher odds of engaging in AMR club activities than second year students and the result was statistically significant(cOR: 3.12, CI= 1.13-8.66, p=0.028).

Having sufficient knowledge about AMR had 2.21 times higher odds of engaging in AMR club activities than having insufficient knowledge and the result was statistically significant (cOR: 2.21, CI:1.10-4.42, p=0.026). Students who were aware about AMR had 3.24 times higher odds of engaging in AMR club activities than those who were not aware and the result was statistically significant(cOR:3.24, CI:1.17-8.96, p=0.023).

Students engaging in extracurricular activities had 12.55 times higher odds of engaging in AMR club activities than those who were engaging in extracurricular activities and the result was statistically significant(cOR: 12.55, CI: 1.64-96.19, p=0.015).

However, after adjusting for confounders, student’s engagement in extracurricular activities had 14.45 times higher odds of engaging in AMR club activities than those who did not and the result was statistically significant (AOR: 14.45, CI: 1.84-113.52, p=0.011)(**Table 3**)

**Table 3:**
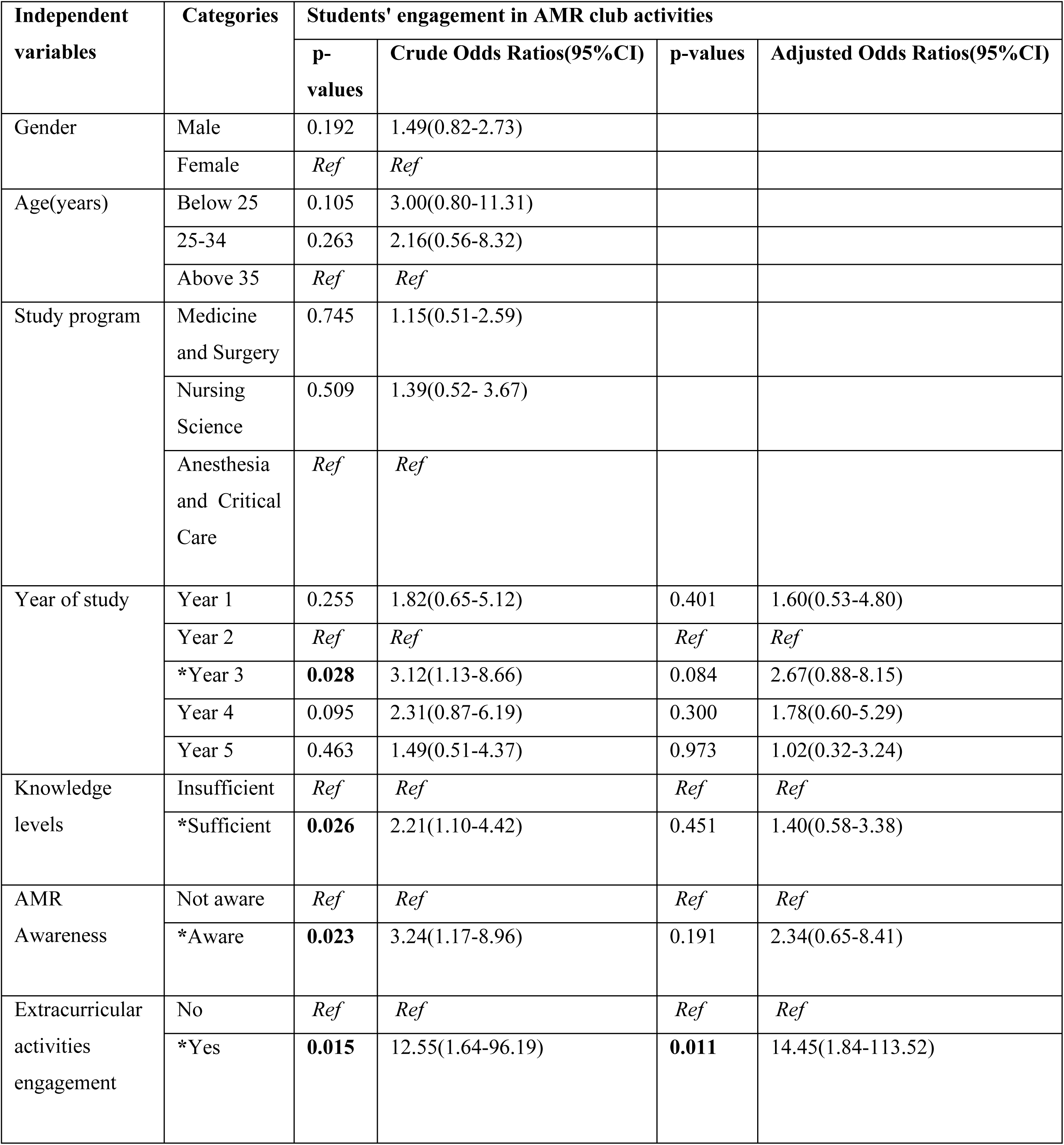
Showing bivariate and multivariable analysis for the factors affecting student engagement in AMR.

| Independent variables | Categories | Students' engagement in AMR club activities |  |  |  |
| --- | --- | --- | --- | --- | --- |
|  |  | p-values | Crude Odds Ratios(95%CI) | p-values | Adjusted Odds Ratios(95%CI) |
| Gender | Male | 0.192 | 1.49(0.82-2.73) |  |  |
|  | Female | *Ref* | *Ref* |  |  |
| Age(years) | Below 25 | 0.105 | 3.00(0.80-11.31) |  |  |
|  | 25-34 | 0.263 | 2.16(0.56-8.32) |  |  |
|  | Above 35 | *Ref* | *Ref* |  |  |
| Study program | Medicine and Surgery | 0.745 | 1.15(0.51-2.59) |  |  |
|  | Nursing Science | 0.509 | 1.39(0.52- 3.67) |  |  |
|  | Anesthesia and Critical Care | *Ref* | *Ref* |  |  |
| Year of study | Year 1 | 0.255 | 1.82(0.65-5.12) | 0.401 | 1.60(0.53-4.80) |
|  | Year 2 | *Ref* | *Ref* | *Ref* | *Ref* |
|  | *Year 3 | **0.028** | 3.12(1.13-8.66) | 0.084 | 2.67(0.88-8.15) |
|  | Year 4 | 0.095 | 2.31(0.87-6.19) | 0.300 | 1.78(0.60-5.29) |
|  | Year 5 | 0.463 | 1.49(0.51-4.37) | 0.973 | 1.02(0.32-3.24) |
| Knowledge levels | Insufficient | *Ref* | *Ref* | *Ref* | *Ref* |
|  | *Sufficient | **0.026** | 2.21(1.10-4.42) | 0.451 | 1.40(0.58-3.38) |
| AMR Awareness | Not aware | *Ref* | *Ref* | *Ref* | *Ref* |
|  | *Aware | **0.023** | 3.24(1.17-8.96) | 0.191 | 2.34(0.65-8.41) |
| Extracurricular activities engagement | No | *Ref* | *Ref* | *Ref* | *Ref* |
|  | *Yes | **0.015** | 12.55(1.64-96.19) | **0.011** | 14.45(1.84-113.52) |

## Discussion

The study findings indicate that a higher proportion of the participants (95.5% of the male students, 82% of the female students) participated in extracurricular activities with students in lower academic years exhibiting lower knowledge levels about antimicrobial resistance compared to their fifth-year counterparts. These findings are consistent with a previous study, which reported higher knowledge levels among clinical students. Additionally, a study in Ghana showed a sequential increase in AMR knowledge across the years of study, suggesting that students were progressively exposed to more AMR concepts as they advanced in their education[10, 15]. Slightly lower levels of AMR knowledge among students have been reported in various studies across Africa For instance, a study in the Democratic Republic of Congo and Ethiopia found that only 55% of students had sufficient knowledge about AMR, while in Zambia, 70% of students had adequate knowledge levels[9, 16, 17]. These studies suggest a significant knowledge gap among medical students about AMR across the continent and attributed to a diverse factors in different countries[10]. Therefore, there is a critical need for improved education on AMR among medical students in Uganda. Incorporating AMR education into the curriculum can ensure a large number of students are well-informed through a sustainable strategy.

The participants demonstrated a positive attitude towards the rational use of antibiotics, with the majority (87%, n= 168) disagreeing that it is appropriate to purchase antibiotics without a prescription. In contrast, a poor attitude was observed among participants in Sudan, where over 59.6% believed it was acceptable to buy medications from a pharmacy without a prescription[18]. Over a half of the participants demonstrated a poor understanding on the spread of drug-resistant infections Less than 50% of the students (n=68, 35.3%) agreed to being at risk of getting an antibiotic-resistant infection as long as they took their antibiotics correctly showcasing little understanding on spread of drug-resistant infections, while less than 83.1% that agreed that antimicrobial resistance affects animal health and production and human health-related science students had a positive feeling that inappropriate use of antimicrobials is one of the reasons for the occurrence of antimicrobial resistance[19]. 83.4% (n=160) agreed that prescribing broad spectrum antimicrobials when narrower spectrum antimicrobials are available increases antimicrobial resistance showcasing high understanding on prescribing practices. More than 90% of the students(n=186, 96.4%) agreed that a strong knowledge of antimicrobials is important in the medical career similar to other studies which shows that the overarching recognition on the need to have strong knowledge about AMR as health professionals enabling them to make better decisions for optimal health care delivery[10].

From the findings, 80% of the students (n=164, 85%) agreed that excessive use of antimicrobials in livestock causes antimicrobial resistance similar to other studies which shows appreciation on the role the one health approach in fight antimicrobial resistance by recognizing other sectors’ importance. About two thirds of the students (n=142, 73.6%) agreed that antimicrobials were overused at the hospital where they had rotated compared to 36% from a previous study[20]. This shows a better attitude towards recognizing misuse as better stewards of antimicrobials. 41.4% of the students (n=80, 41.4%) agreed that new antimicrobials will be developed in the future that will keep up with the problem of resistance showcasing poor understanding on the complexity of AMR, and the multifaceted approach required.

Students’ engagement in extracurricular activities was associated with engagement in AMR club activities even after adjusting for other factors because since AMR club was an extracurricular activity alongside the others, it was easier for the students already engaged in the other activities to take on AMR club activities.

Most students engaged in AMR club activities due to influence from peers, university support and mentors while the least engaged to get information and learning. This is mainly because students can easily access their peers and are more inspired and influenced by the actions of people within the same bracket[21].

## Conclusion

In conclusion, the study reports a sequential increase in knowledge levels on AMR across the years of study amongst the study participants. The study showed that healthcare students appreciated the need and importance of AMR training through a One Health Approach and the importance of good infection prevention and control measures in addressing AMR. Influence from peer, university support and inspiration by peer mentors work were found to be the major reasons behind students’ engagement in AMR related extracurricular activities.

## Recommendations

From our findings, we recommend that, an AMR and AMS course unit should be incorporated into the preservice training of healthcare students so that there is a greater understanding of Antimicrobial Resistance and the topic and potential mitigation efforts. Furthermore, there is need to emphasize on the value of extracurricular engagement in AMR among healthcare students and to leverage on peer support and mentorship to promote holistic learning and effective community engagement in AMR. Lastly, we recommend a broader study incorporating multiple institutions in investigating the drivers and barriers of healthcare students in extracurricular AMR engagements to obtain generalizable results that can inform the country’s AMR policies.

## Limitations

The study was done in one university therefore it may be difficult to generalize the findings at a country level.

## Data Availability

All relevant data are within the manuscript and its Supporting Information files

## Acknowledgements

The authors thank all the students who participated in the study for their feedback. We are grateful the Foundation to Prevent Antibiotic Resistance for grant awarded to support this project that was aimed at catalyzing grassroot engagement of tertiary level students in different countries in Africa. We thank Busitema University Antimicrobial Resistance and Stewardship Club and Busitema University Research and Innovation Association for allowing us to use their structures to implement the study. We thank ReAct (Action on Antibiotic Resistance) Africa and Zihi Institute for the technical support in implementation of this study.

